# Performance of RAPID noncontrast CT stroke platform in large vessel occlusion and intracranial hemorrhage detection

**DOI:** 10.1101/2023.08.11.23294007

**Authors:** Vivek Yedavalli, Jeremy J Heit, Seena Dehkharghani, Hafez Haerian, John M. Mcmenamy, Justin Honce, Vincent M Timpone, Christopher Harnain, Andrew Kesselman, Anthony Filly, Adam Beardsley, Brian Sakamoto, Christopher Song, James L Montuori, Benjamin Navot, Francisco Villar Mena, Dan-Victor Giurgiutiu, Felipe Kitamura, Fabricio Oliveira Lima, Henrique Coelho Silva, Francisco José Arruda Mont’Alverne, Gregory W Albers

**Author notes:** **Corresponding Author:** Vivek Yedavalli, MD, MS, 600 N. Wolfe St., Phipps B112-D, Baltimore, MD 21287.

## Abstract

**Background:** Noncontrast CT (NCCT) is used to evaluate for intracerebral hemorrhage (ICH) and ischemia in acute ischemic stroke (AIS). Large vessel occlusions (LVOs) are a major cause of AIS, but challenging to detect on NCCT.

**Aims:** The purpose of this study is to evaluate an AI software called RAPID NCCT Stroke (RAPID, iSchemaView, Menlo Park, CA) for ICH and LVO detection compared to expert readers.

**Methods:** In this IRB approved retrospective, multicenter study, stand-alone performance of the software was assessed based on the consensus of 3 neuroradiologists and sensitivity and specificity were determined. The platform’s performance was then compared to interpretation by readers comprised of eight general radiologists (GR) and three neuroradiologists (NR) in detecting ICH and hyperdense vessel sign (HVS) indicating LVO. Receiver operating characteristics (ROC) curve was used to evaluate the performance of each reader. P < 0.05 was considered significant.

**Results:** A total of 244 cases were included. Of the 244, 115 were LVOs and 26 were ICHs. One hundred three cases did not have LVO nor ICH. Stand-alone performance of the software demonstrated sensitivities and specificities of 96.2% and 99.5% for ICH and 63.5% and 95.1% for LVO detection.

Compared to all 11 readers and eight GR readers only respectively, the software demonstrated superiority, achieving significantly higher sensitivities (63.5% versus 43.6%, p < 0.0001 and 63.5% versus 40.9%, p =0.001).

**Conclusion:** The RAPID NCCT Stroke platform demonstrates superior performance to radiologists for detecting LVO from a NCCT. Use of this software platform could lead to earlier LVO detection and expedited transfer of these patients to a thrombectomy capable center.

## Introduction

Acute stroke secondary to ischemia or hemorrhage is among the leading causes of death and disability worldwide^1^. Acute ischemic stroke (AIS) comprises the majority of acute strokes (85-90%) ^2^. Despite accounting for a small fraction of acute stroke, intracerebral hemorrhage (ICH) confers a high risk of mortality^2^.

Neuroimaging is crucial to contemporary stroke management paradigms. Noncontrast CT (NCCT) is the most commonly used imaging modality to screen for intracranial hemorrhage (ICH) and assess for early ischemic changes^3^. Large vessel occlusions (LVOs) may also be detectable on NCCT through the identification presence of a hyperdense vessel sign (HVS), a variably present feature of intracranial LVO^4^. NCCT has a high sensitivity for detecting early ICH^2^ but has a sensitivity of only approximately 40% for detecting early signs of ischemic stroke^5^. Sensitivity is also roughly 50% for detecting HVS as a marker of LVO^4^. In order to facilitate management in the hyperacute setting, artificial intelligence (AI) methods are now increasingly used with NCCT to detect both ICH and/or AIS^6^ and may serve as valuable adjuncts for radiologic evaluation.

Many comprehensive stroke centers perform both a NCCT and CTA in patients with a suspected stroke. However, community hospitals frequently obtain only an NCCT scan initially and then discuss the case with a neurologist and/or radiologist then subsequently obtain a CTA in selected cases. This approach can lead to significant delays in LVO diagnosis and lengthen the time to transfer the patient to a thrombectomy center. AI software has the potential to immediately evaluate a NCCT and notify the treating physicians that an LVO is suspected, which could lead to substantial improvements in workflow.

## Aims and Hypotheses

RAPID NCCT Stroke (iSchemaView, Menlo Park, CA; RAPID) is a multi-module fully automated AI platform developed to detect both ICH and LVO (defined as occlusion of the middle cerebral artery (MCA) and/or distal internal carotid artery (ICA)) by integrating multiple findings available from the NCCT. We assessed the sensitivity and specificity of this software for detection of ICH and LVO as a) a stand-alone platform, and b) as compared to general radiologists and neuroradiologists. We hypothesize that the AI software outperforms general radiologists (GR) and is non-inferior to neuroradiologists (NR) in a multicenter retrospective evaluation of acutely presenting patients with suspected stroke.

## Methods

### Subjects

This retrospective, multicenter study was conducted in compliance with the Health Insurance Portability and Accountability Act (HIPAA) and was approved by the institutional review boards (IRB; Advarra central IRB Pro00049230). Cases were obtained from consecutive emergency room (ER) scans obtained in patients suspected of having an acute stroke in two of the centers and from research studies that enrolled acute LVO and ICH patients. In total, 244 cases were included in the analysis.

### Definitions of pathology

ICH was defined as any type of intracranial hemorrhage including intraparenchymal, subdural, epidural, subarachnoid, and intraventricular hemorrhages^5^. LVO was defined as distal intracranial ICA and M1 segment of the MCA^7^. Lastly, HVS was determined to be a segmental hyperdensity which is comparatively more hyperdense with respect to the contralateral hemisphere and corresponding to the expected location of the distal intracranial ICA and/or M1 segment of the MCA^4^.

### Scanners

This study is composed of cases from different CT vendors. In total, 93 cases were obtained from Siemens scanners (Siemens Healthineers, Erlangen, Germany), 50 from GE Healthcare (GE Healthcare, Wauwatosa, WI, USA) 44 from Phillps (Koninklijke Philips, Amsterdam, Netherlands), 57 from Toshiba (Toshiba, Minato City, Japan).

### RAPID NCCT Stroke Development

NCCT imaging data were anonymized and translated into a spatial 3D model.This AI software uses neural networks and automated segmentation techniques based on predefined thresholds for identification of ICH, HVS, and Alberta Stroke Program Early CT Score (ASPECTS).

A proprietary algorithm was used to determine if an LVO is likely to be present based on a combination of features derived from the assessment of HVS as well as the specific regions of involvement on ASPECTS modules. The specific modules that are used in RAPID NCCT are RAPID ICH 3.0, RAPID ASPECTS 3.0 and RAPID HVS.

Eleven total board-certified GR and NR readers assessed all 244 NCCT scans using local installations of DICOM images through a viewing platform (Osirix, Geneva, Switzerland). Expert readers interpreted the images using only soft tissue kernels with 5 mm slice thickness, including multiplanar reformats. All readers performed interpretations blinded to the AI software results and clinical information. Readers then assessed for ICH, and if ICH was not present, they then assessed for suspected LVO. The reference truth for ICH was based on a consensus of two of three neuroradiologists evaluating the NCCT scan using the same parameters. Scans that expert readers identified with ICH were classified as No LVO for LVO performance assessment.

Stand-alone performance was based on the reference LVO assessment which was determined by a consensus of two of three neuroradiologists based on a CTA performed concurrently with the NCCT. CTA assessments were performed on maximum intensity projection (MIP) images based on 3 mm slice thickness using a soft tissue kernel six weeks after the initial NCCT assessment.

A board-certified NR (JJH, 10 years of experience) independently reviewed cases deemed positive for LVO to screen for suspected vessel calcification in the distal intracranial ICA or M1 segment of the MCA. This review was based on binary determination of presence or absence of vessel calcification. Cases interpreted as positive for ICH were not assessed for vessel calcifications and classified as ‘No LVO.’

The primary endpoint was sensitivity and specificity of the software as compared to those of the GR and NR readers who were blinded to the CTA results. In total, eight GR and three NR expert readers (different individuals than the experts who determined the reference standard on the CTAs) participated in this phase of the study. Readers were instructed to consider both the presence of HVS as well as early parenchymal signs of brain ischemia when making their determination of LVO. The primary hypothesis was that the automated software would have a higher sensitivity than GR for detecting LVO and be non-inferior to NR readers. If non-inferiority was achieved, then the software would be tested for superiority against all readers as well as NR readers alone. Overall accuracy was also compared between the software and individual readers to assess for both sensitivity and specificity.

### Statistical analysis

Sensitivity and specificity analyses were calculated by comparison of the software results to the CTA reference standard for LVO for the stand-alone evaluation. Subsequently, the sensitivity and specificity of the software was compared to the GR and NR readers. Using the ratings (1 to 5) scale that readers used to express their confidence in their LVO call, the readers’ ROC curves were then compared to the overall ROC curves.

## Results

Ultimately, a total of 244 cases were included in this study. Of the 244, 115 were LVO (115/244, 47.1%) and 26 (26/244, 10.7%) were ICHs based on the consensus of two of three expert neuroradiologists. One hundred three cases (103/244, 42.2%) were independently reviewed for suspected vessel calcification after initial screening. One hundred three cases (103/244, 42.2%) did not have LVO nor ICH.

### NCCT Stroke stand-alone performance

LVO: The software identified 73 true suspected LVOs (73/115, 63.5%) with 42 false negatives (42/115, 36.5%) It also correctly categorized 90 cases (90/115, 78.2%) where LVO was not present with five false positives (5/115, 4.3%). This resulted in a sensitivity of 63.5% (95% CI:54.4% - 71.7%) and specificity of 95.1% (95% CI: 89.1% - 97.9%).

ICH: The software identified suspected ICH in 25 of the 26 cases (25/26, 96.1%). Among the 217 ICH-negative cases, the software correctly identified the absence of ICH in 216 (216/217, 99.5%), with one false positive (1217, 0.4%). The overall sensitivity and specificity for ICH detection were 96.2% (95% CI: 81.1% - 99.3%) and 99.5% (95% CI: 97.4% - 99.9%), respectively.

### Comparison with GR and NR readers - LVO detection

Eleven total readers independently assessed the presence of LVO. Sensitivities ranged from 20% up to 64.3% with specificities ranging from 70.9% up to 100%. See figure 1 for details of the independent readers in comparison to the platform. Comparison of sensitivities of RAPID and expert readers are shown in Table 1.

**Table 1.** Sensitivity comparison of GR and NR readers.

| <u>Reader Description</u> | <u>N Readers</u> | <u>RAPID</u><br><u>Sensitivity</u> | <u>Reader Sensitivity</u> | <u>SE</u><br><u>(Readers)</u> | <u>P-value</u> |
| --- | --- | --- | --- | --- | --- |
| Average of all general radiologists | 8 | 0.64 | 0.41 | 0.065 | 0.001 |
| Average of all Readers | 11 | 0.64 | 0.44 | 0.055 | <.0001 |
| Average of neuroradiologists | 3 | 0.64 | 0.51 | 0.061 | 0.008 |

**Figure 1a:**
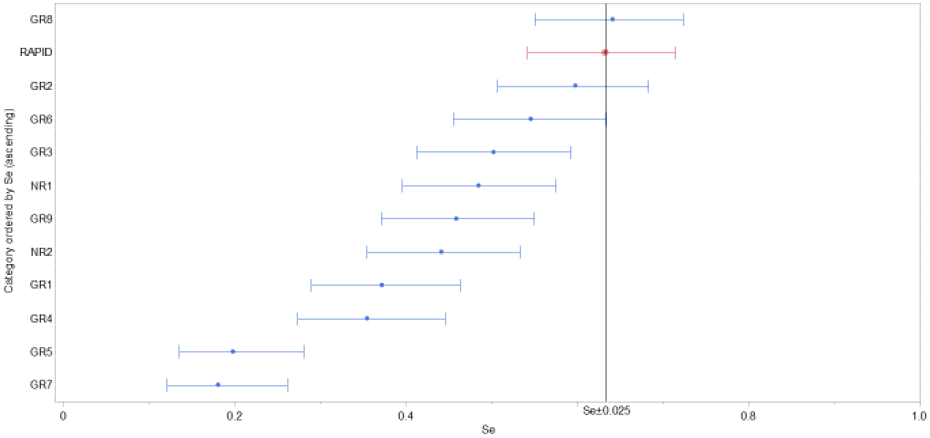
Comparison of the sensitivities with 95% for each reader separated into general and neuroradiologists (blue) and that of RAPID (red). RAPID demonstrates sensitivity higher than all expert readers except one.

**Figure 1b:**
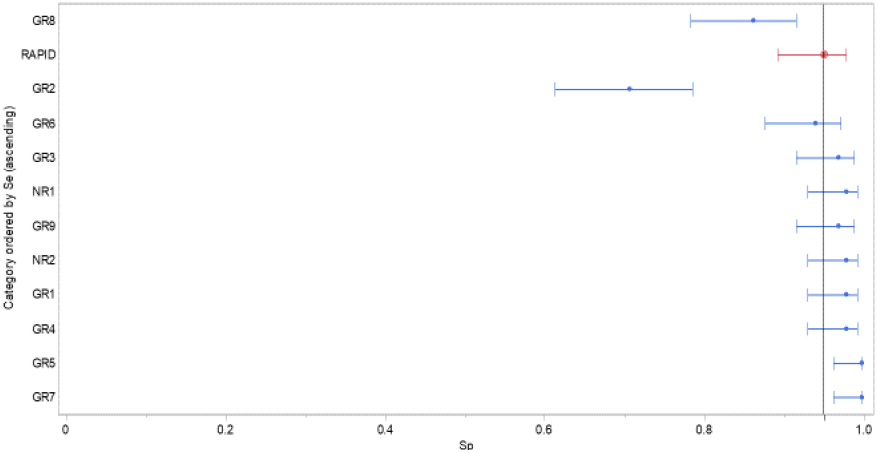
Comparison of the specificities with 95% for each reader separated into general and neuroradiologists (blue) and that of RAPID (red). RAPID demonstrates a specificity comparable to that of expert readers.

### Diagnostic performance of GR and NR readers - LVO detection

ROC analyses measuring diagnostic performance of the readers revealed a wide range in AUC from 0.592 (GR7) to 0.823 (GR3). See figure 2.

**Figure 2:**
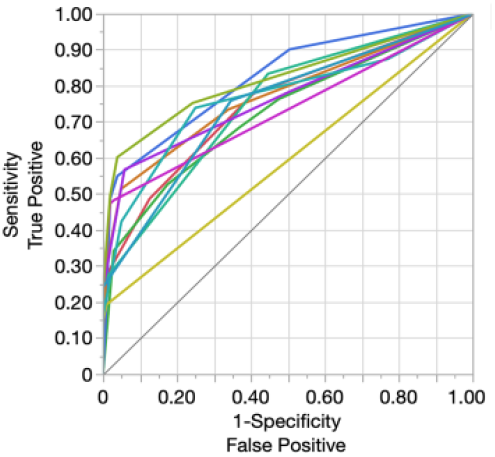
ROC curves for individual readers. Using the ratings (1 to 5) scale that readers used to express their confidence in their LVO call we compare reader ROC curves and the overall ROC curves. RAPID uses a decision tree to arrive at classification, there is not an underlying score to generate a ROC curve for RAPID.

### Non-inferiority/superiority testing

The primary endpoint was to compare the stand-alone performance of the RAPID NCCT platform to the 11 expert GR and NR readers. We hypothesized that the RAPID NCCT Stroke platform would demonstrate superior performance to the eight GR readers and non-inferior performance to the three NR readers. In assessing non-inferiority of the platform compared to the 11 readers, the platform achieved both non-inferiority as well as superiority, with a significantly higher sensitivity (63.5% versus 43.6%, p < 0.0001).When compared to the eight GR readers only, the platform also showed both non-inferiority and superiority with significantly higher sensitivity (63.5% versus 40.9%, p = 0.001). Non-inferiority and superiority were also established for the comparison with the 3 NR readers alone. Please refer to table 1 for additional detail. The overall agreement with the reference standard was also highest with the software.

### Vessel calcification assessment

Of the 244 cases screened, a total of 103 cases were classified as suspected vessel calcifications in the horizontal segment of the MCA. Two cases showed calcification and were classified as true negatives (2/103, 1.9%). An additional 16 true negative cases had calcifications noted elsewhere (16/103, 15.5%).

## Discussion

NCCT is a readily available and efficient imaging modality for excluding ICH and detecting early signs of ischemia but CTA is typically required for confirming large vessel occlusions. In this investigation, we focus on the utility of NCCT as it pertains to detecting LVOs and ICH. We report that the RAPID NCCT module has a superior sensitivity for detecting LVO compared with general radiologists and neuroradiologists. The platform also showed excellent sensitivity and specificity in identifying ICH. Our findings suggest that this software has potential utility as an adjunct for neurologists, neurointerventionalists, and radiologists in routine clinical practice for increasing the accuracy of LVO and ICH detection on NCCT.

Community hospitals often perform an initial NCCT in patients who present with new neurological symptoms. Subsequently, CTA may be performed if the initial CT does not disclose the diagnosis or if an LVO is suspected. However, CTA acquisition can be challenging in smaller community hospitals and rural centers across the world because of the need for iodinated contrast, concerns for increased radiation exposure, and technical expertise needed to obtain a study that is considered diagnostic^8^. Furthermore, the delay between clinical assessment/evaluation of the NCCT and when the CTA is obtained can be considerable, often >30 minutes, for the aforementioned reasons. AI software has the potential to address these concerns by substantially reducing this delay when providing an immediate notification that an LVO is suspected. This notification could expedite additional imaging or urgent transfer to a thrombectomy capable center.

NCCT is the most common screening modality when stroke is suspected but excluding ICH is essential in order to potentially administer thrombolysis. NCCT is highly sensitive and specific for detecting ICH of all subtypes^9^. For instance, a meta-analysis by Dubosh et al found NCCT within six hours of symptom onset has a sensitivity and specificity of 98.7% and 99.9% for detection of spontaneous subarachnoid ICH^10^. The high specificity and sensitivity of NCCT makes it the primary screening modality for not only subarachnoid ICH but all forms of ICH^11^.

Several prior studies have utilized AI platforms in detecting ICH. Goyal used machine learning (ML) techniques in detecting ICH on NCCT with sensitivities ranging from 95-100% and specificities between 85-100%^12^. Seyam et al similarly used a developed AI based software geared toward ICH detection on NCCT with a sensitivity of 87.2% and specificity of 93.9%^13^. Others have also utilized deep learning (DL) based models in ICH detection on NCCT with similarly strong results^14–16^. Software performance for detecting ICH is similar to the Goyal et al and exceeds that of several aforementioned reports, demonstrating a sensitivity of 96.2% and specificity of 99.5%. The Rapid ICH 3 module that is used in the NCCT stroke platform has a sensitivity of 97% and a specificity of 100% for detecting ICHs of any subtype of 0.4 ml or larger^17^ .

When compared to ICH detection, detecting LVO is more challenging on NCCT. The HVS is an important sign of LVO on NCCT evaluation. However, prior studies have reported sensitivities ranging from 17-52% with specificities approaching 100% for HVS detection ^3,4,18^. Despite the variability in detection, the high specificity of HVS makes it particularly useful in early LVO detection for expeditious transfer to a larger center and/or emergent treatment.

Similarly, to ICH evaluation, LVO detection on NCCT using AI applications is a growing area of interest. A systematic review by Shlobin et al concluded that AI applications for LVO detection on CT imaging have reasonable accuracy and show promise as an adjunct tool in the decision-making process^19^. More recently, a study by Olive-Gadea et al developed a DL technique that identified LVOs on NCCT with 83% sensitivity and 71-85% specificity with human interpretation as the ground truth ^20^. However, concerns with this algorithm were raised based on the cohort which the algorithm was applied to, the use of clinical and imaging data (NIHSS in addition to NCCT) and the generalizability. The generalizability concern was mainly based on the remote hospital setting the original study performed as the quality of NIHSS assessments may be higher at larger academic institutions ^21^. In our study, the software detected LVO with a sensitivity of 63.5% and a specificity of 95.1% using only NCCT data. The higher specificity can be particularly useful in smaller centers where mobilizing resources to transfer patients to comprehensive stroke centers may be more challenging.

As with previous reports, the software performance was compared to human expert interpreters. We hypothesized that the platform will be superior to that of GR readers and non-inferior to NR readers. Eleven GR and NR readers independently interpreted the cases, showing a sensitivity range of 20-64.3% and specificity of 70.9-100%. When compared to only the eight GR readers, the platform demonstrated superior performance sensitivities (63.5% versus 40.9%, p = 0.001). Furthermore, with the inclusion of NR readers, the platform’s performance was not only non-inferior but still superior with respect to sensitivity (63.5% versus 43.6%, p < 0.001).

Our study has several limitations to acknowledge. Firstly, it is limited due to the retrospective design. It is nevertheless strengthened by the robust sample size acquired from five centers that utilize different CT vendors, thus improving generalizability. Secondly, the HVS can be difficult to differentiate from calcifications.

We identified cases where calcifications may also be present, all of which were confirmed as true negatives after expert interpretation in order to address this potential limitation. Lastly, although the sensitivity of the platform for independently detecting LVO in absolute measures may be considered low, it is a significant improvement compared to current practice (64% versus 43% for combined GR and NR expert readers). Nevertheless, this study lays the foundation for future investigations exploring the combined sensitivity of the platform with the assessments of interpreting physicians, including neurologists, neurointerventionalists, and radiologists for improved LVO detection. Future studies building upon the current results will also be necessary for detecting posterior circulation and medium vessel occlusions.

## Conclusion

The RAPID NCCT Stroke platform demonstrated superior performance to GR and NR readers, suggesting that this software can function as a useful adjunct tool for stroke physicians for expediting the detection of LVOs and urgent transfer to a thrombectomy capable center. Prospective studies are needed for further validation.

## Data Availability

Data can be made available upon reasonable request.

## Acknowledgements

None

## Author Contributions

Vivek Yedavalli, analysis and interpretation of data and drafting the manuscript. Greg Albers: design and conceptualization of the study and drafting the manuscript All authors: editing and reviewing.

## Declaration of Interest

Drs. Jeremy Heit and Vivek Yedavalli disclose roles as consultants for RAPID (iSchemaView, Menlo Park, CA). Dr. Greg Albers is the co-founder of RAPID.

## Funding

No funding source for this investigation.

## Notes

### Author Declarations

This retrospective, multicenter study was conducted in compliance with the Health Insurance Portability and Accountability Act (HIPAA) and was approved by the institutional review boards (IRB; Advarra central IRB Pro00049230).

